# Airway Mucus Plugs in Community-Living Adults: A Study Protocol

**DOI:** 10.1101/2024.05.15.24307439

**Authors:** Maya Abdalla, Rim Elalami, Michael H Cho, George T O’Connor, Mary Rice, Michael Horowitz, Neda Akhoundi, Andrew Yen, Ravi Kalhan, Alejandro A. Diaz

**Author notes:** **Corresponding Author:** Alejandro A. Diaz, MD, MPH, Division of Pulmonary and Critical Care, Department of Medicine, Brigham and Women’s Hospital 75 Francis St, Boston, MA 02115.

## Abstract

**Introduction:** Mucus pathology plays a critical role in airway diseases like chronic bronchitis (CB) and chronic obstructive pulmonary disease (COPD). Up to 32% of community-living persons report clinical manifestations of mucus pathology (e.g., cough and sputum production). However, airway mucus pathology has not been systematically studied in community-living individuals. In this study, we will use an objective, reproducible assessment of mucus pathology on chest computed tomography (CT) scans from community-living individuals participating in the Coronary Artery Risk Development in Young Adults (CARDIA) and Framingham Heart Study (FHS) cohorts.

**Methods and analysis:** We will determine the clinical relevance of CT-based mucus plugs and modifiable and genetic risk and protective factors associated with this process. We will evaluate the associations of mucus plugs with lung function, respiratory symptoms, and chronic bronchitis and examine whether 5-yr. persistent CT-based mucus plugs are associated with the decline in FEV_1_ and future COPD. Also, we will assess whether modifiable factors, including air pollution and marijuana smoking are associated with increased odds of CT-based mucus plugs and whether cardiorespiratory fitness is related in an opposing manner. Finally, we will determine genetic resilience/susceptibility to mucus pathology. We will use CT data from the FHS and CARDIA cohorts and genome-wide sequencing data from the TOPMed initiative to identify common and rare variants associated with CT-based mucus plugging.

**Ethics and Dissemination:** The Mass General Brigham Institutional Review Board approved the study. Findings will be disseminated through peer-reviewed journals and at professional conferences.

**Strengths and limitations of this study:**

- Utilization of data from two well-characterized large community-based US cohorts.
- Use of chest CT scans to identify and quantify mucus plugs, providing a more objective and reproducible measure of airway pathology.
- Use of whole-genome sequencing to identify common and rare genetic variants associated with mucus pathology.
- Only the inclusion of participants self-identified as non-Hispanic white and non-Hispanic black.
- A limitation of retrospective study design using prospectively collected data.

## Introduction and Rationale

Mucus pathology plays a critical role in airway diseases, such as chronic bronchitis (CB), asthma, and chronic obstructive pulmonary disease (COPD) [1–9]. COPD affects ∼29 million people and is the 4th leading cause of death in the US [10, 11]. Mucus pathology and plug formation are complex processes in response to multiple inhaled noxious particles, including cigarette smoke and air pollution [1, 12, 13]. The public health and clinical relevance of airway mucus pathology are better understood through its clinical manifestations, including chronic cough and phlegm and chronic mucus hypersecretion/CB [14–19]. The prevalence of chronic respiratory symptoms in primary care is up to ∼40% [20]. In a large study, 32% of ∼98,000 community-living individuals with normal lung function reported chronic respiratory symptoms. More importantly, those symptoms were indicators of an increased risk of future hospitalization and mortality [14].

We and others have also demonstrated that in smokers with COPD, mucus plugs occluding the lumen of medium-to-large-sized airways (i.e., 2-10-mm-lumen diameter), those visualizable on CT, are associated with impaired lung function, worse quality of life, and increased all-cause mortality [6, 7, 21]. We posit that CT assessment of airway mucus pathology applies to community-living individuals [22, 23].

Extensive research has been conducted on chronic respiratory disorders. While this has brought new insights into diseases’ clinical variability and biological basis, attention has been limited to patients with established and even advanced disorders. Suppose clinical care is to identify early risk factors and prevent disease. In that case, new work is needed to identify those at high risk of future diseases, those who may have poor lung health but do not yet meet existing diagnostic criteria for an established airway disease [17]. We have shown that smokers have persistent mucus plugs on CT over a 5-year follow-up [6], but whether this pattern represents a risk factor for airway disease in community-living adults is unknown.

In this study, we will also determine the role of modifiable factors, such as air pollution, marijuana, and fitness, on mucus pathology, which may inform specific preventive and treatment strategies for lung health (e.g., air purification). Exposure to air pollution and cigarette smoke induce pathologic mucus production in the airways; however, not all individuals exposed to the same set of factors develop comparable mucus dysfunction and its consequences, suggesting genetic susceptibility or resilience may have a role [23].

Our team has implicated TP53 gene variants in increased mucus synthesis and sustained the lifespan of metaplastic bronchial mucous cells. Further, smokers with the p53^Arg^ are at increased risk of CB (OR=1.59; p=0.03) [24].

We propose that CT-based mucus plugs are an intermediate phenotype that will enable us to investigate the clinical implications of mucus pathology and assess its contribution to the risk of airway disease. To test our hypothesis, we will use data from two population-based cohort studies: The Coronary Artery Risk Development in Young Adults (CARDIA) study [25] and the Framingham Heart Study (FHS) [26, 27]. The study protocol presented here was funded by the National Heart Lung and Blood Institute (Grant R01-HL164824).

### Specific Aims

Our overall hypothesis is that CT-based mucus plugs are an intermediate phenotype that will enable us to examine the clinical implications of mucus pathology and gauge its contribution to airway disease risk in population-based studies. The study has three specific aims:

1. Determine whether CT-based mucus plugs are associated with reduced FEV_1_, respiratory symptoms, and CB in community-living adults.
2. Determine whether modifiable exposures (long-term air pollution, marijuana use, and fitness) are associated with CT-based mucus plugs.
3. Assess the genetic resilience/susceptibility to mucus pathology by examining whether common and rare variants are associated with CT-based mucus plugging in community-living adults.

We will use existing imaging, clinical, environmental, and genetics data already collected from the CARDIA and FHS cohort, thus not exerting additional burden to the study participants and avoiding further radiation exposure.

## Methods and Analysis

### Study design

The present will use prospectively collected data from the CARDIA and FHS cohorts. CARDIA is a United States (US) National Institutes of Health (NIH)-funded population-based cohort of 5,115 young adults recruited from 1985 to 1986 to explore the evolution of cardiovascular disease risk factors [25]. Participants were examined at four sites across the US during enrollment. In-person examinations occurred at baseline (year [Y] 0), Y2, Y5, Y7, Y10, Y15, Y20, Y25, and Y30. Examinations collected demographic, anthropometric, clinical (e.g., symptoms and tobacco and marihuana smoking history), fitness, chest CT, and spirometry data in a standardized manner. CARDIA also took and kept blood samples. Participants’ blood samples were sent to the NIH Trans-Omics for Precision Medicine (TopMed) program to perform whole-genome sequencing. We will use chest CT images from the Y20 (n = 3,122) and Y25 (n = 3,498) examinations to score airway mucus plugs.

The FHS started in 1948 as a prospective investigation of cardiovascular disease, and we will use participants from the FHS Multidetector Computed Tomography 2 (MDCT2) sub-study (n=2750). Participants of the MDCT2 sub-study are from the FHS Offspring Cohort (1971) and the FHS Third Generation (2002) cohort [27]. The FHS Third Generation participants are the children of the Offsprings. FHS also collected demographic, anthropometric, clinical (e.g., symptoms and tobacco smoking history), air pollution, spirometry, and chest CT data in a standardized manner. FHS participants’ blood samples were also sent to the NIH TopMed program to perform whole-genome sequencing. We will use chest CT images from the MDCT2 sub-study to identify and score airway mucus plugs. Characteristics of the Y20 CARDIA and FHS participants are depicted in Table 1.

**Table 1.** Characteristics of Year 20 CARDIA and FHS participants.

| **Characteristic** | **CARDIA**<br>**(n=3,122)** | **FHS**<br>**(n=2,750)** |
| --- | --- | --- |
| Age, years | 45 (4) | 59 (12) |
| Female sex* | 57% | 50% |
| White race* | 53% | 100% |
| Education status |  |  |
| High school or less | 23% | 23% |
| More than high school | 28% | 31% |
| College degree or more | 50% | 46% |
| BMI, kg/m<sup>2</sup> | 30.2 (7.2) | 28.5 (5.4) |
| Tobacco smoking status |  |  |
| Never | 62% | 48% |
| Current | 18% | 6% |
| Former | 20% | 45% |
| Pack-years smoked | 5.1 (9.4) | 9.6 (16.1) |
| Lifetime marihuana smoking,<br>No. of joints/years | 366/20<br>yrs. | - |
| PM<sub>2.5</sub>, mcg/m<sup>3</sup> | - | 8.9 (1.5) |
| Fitness (treadmill duration), min | 7.1 (2.7) | - |
| Cough | 9% | 7% |
| Phlegm | 9% | 6% |
| Dyspnea | 19% | 11% |
| Asthma | 13% | 6% |
| Episodes of bronchitis<sup>^</sup>/CB | 4% | 7% |
| Spirometric COPD | 8% | 21% |
| FEV<sub>1</sub> % predicted | 92 (16) | 98 (15) |
| FVC % predicted | 94 (15) | 102 (13) |
| FEV<sub>1</sub>/FVC | 0.77 (0.1) | 0.74 (0.1) |
| Emphysema<sup>&</sup>/LAA-950, % on CT | 2.4% | 26.9 (7.2) |
| Airway wall thickness, mm | - | 1.1 (0.1) |

Data are presented as mean (standard deviation) and proportion, %. Percentages were calculated with data available for each variable. \*Race and sex data are from Y25 exam. ^CB questions are available for FHS only. Emphysema (yes/no) was visually assessed (CARDIA). LAA, % of lung attenuation areas under -950 Hounsfield units. Cells with – means no data.

### Patient and Public Involvement Statement

Patients and/or the public were not involved in the design, conduct, reporting or dissemination plans of this research.

### Eligibility

In the CARDIA cohort, eligibility was limited to young adults aged 18-30 years in 1985 to 1986 with no evidence of long-term diseases or disabilities. In the FHS cohort, eligibility was limited to participants aged 20 years or older and had at least one parent who was a member of the Offspring Cohort [27]. All individuals known to be eligible for the inclusion in the Third Generation Cohort were sent invitation letters along with response letters, and high interest was indicated by return of response cards, which led to more participation than anticipated.

### Participant Background

At Y20, the mean age was 45 years; 38% were smokers, and 9% - 19% reported cough, phlegm, or dyspnea. CARDIA included a balanced number of male and female and Black and White participants, whereas FHS had well-balanced sexes (M/F) but only included White participants. In the FHS-MDCT2 cohort, the mean age was 59 years; 51% were former and current smokers, and 7-11% reported respiratory symptoms. Participants of both population-based cohort studies include varying cardiorespiratory fitness, air pollution exposure, and tobacco and marijuana smoking histories.

### Study Data Collection

The CARDIA and FHS studies used standardized questionnaires and procedures to collect imaging, clinical (e.g., marijuana history), environmental, fitness, and genetic data. Also, the methodology to assess mucus plugs on CT scans for this study is detailed below. The Institutional Review Board of Mass General Brigham approved this study. All participants provided written consent.

### Chest CT scans

All participants underwent chest CT scanning. CARDIA patients underwent a noncontrast electrocardiography-gated chest CT scan reconstructed at 2.5 to 3 mm slice thickness [28]. FHS-MDCT2 sub-study patients underwent lung volumetric CT scans in the supine position, reconstructed by a sharp lung algorithm with a 0.625 mm thickness and a 50 cm field of view [29]. Although there are differences in reconstructions between cohorts, the quality of CT scans reconstruction parameters is adequate for this study to survey the airways.

### Defining Mucus Plugs

A mucus plug is defined as an opacity that completely occludes the lumen of an airway on sequential slices, regardless of the generation and orientation of the bronchus (Figure 1) [3,4]. The opacity can be round or cylindrical in shape and found in one or many airway branches. Plugging is identified in medium-to-large-sized airways on CT.

**Figure 1.**
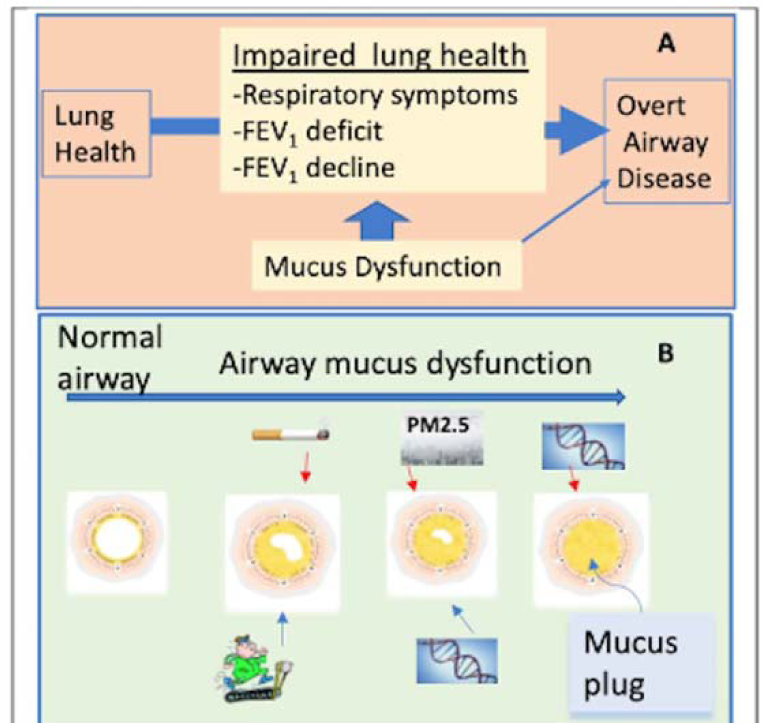
**A. Progression from healthy to overt airway disease.** Impaired lung health is between these extremes. Mucus dysfunction is associated with impaired lung heath through multiple factors yet provides a target to intercept pulmonary diseases before they become “chronic.” **B. Airway mucus dysfunction.** Tobacco, marijuana, and PM2.5 increases mucus plugs whereas fitness can protect against plug formation. Genetic variants resilience or susceptibility may also play a role in airway mucus biology.

### Mucus Plug Score

The scoring method was developed based on the number of pulmonary segments with mucus plugs regardless of the orientation of the bronchus (Figure 2). For instance, a mucus plug identified in the lower right lobe’s posterior and lateral basal segments yields a score of two. The score will range from 0 to 18, where 0 means no plugging and higher numbers indicate a greater extent of plugs.

**Figure 2.**
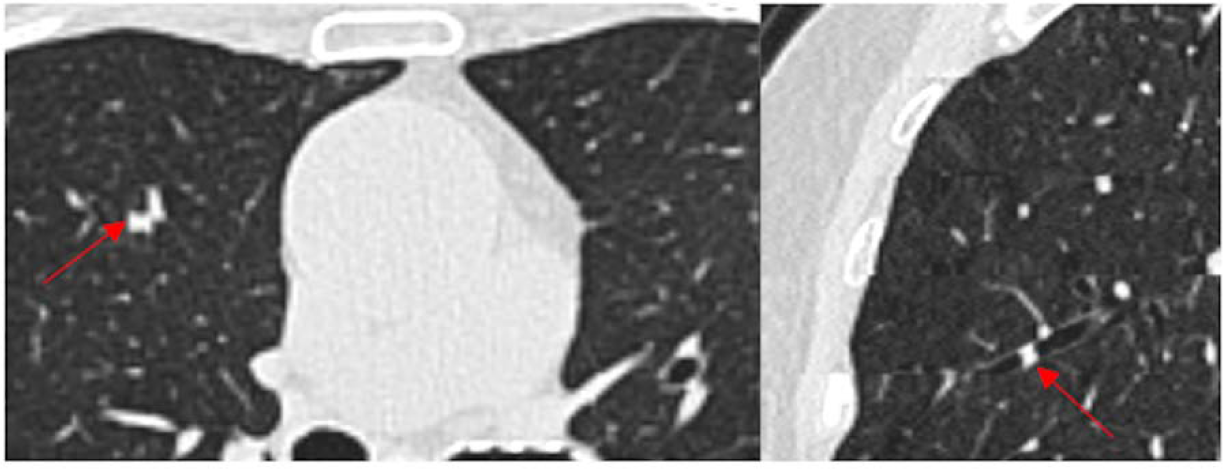
**A** Axial CT image showing a mucus plug occluding upper lobe. **B** The same mucus plug in the sagittal plane.

### Reading Method

The readers who will score mucus plugs comprise thoracic radiologists, a pulmonologist, physicians, and graduate students. Participating readers undergo training, which consisted of identifying a variety of mucus plugs on a set of typical CT images, scoring a training set of CT scans to learn the scoring system, and scoring a sample set of 20 CT scans to verify the inter-reader agreement. The concordance correlation for inter-reader agreement of mucus plug score previously reported was 0.84 (95% Cl 0.81-0.88) [6]. The Applied Chest Imaging Laboratory also has developed an automated method to detect mucus plugs, providing an alternate way to assess airway mucus pathology on CT images.

### Spirometry

Spirometry protocols were designed following the American Thoracic Society (ATS)/European Respiratory Society (ERS) guidelines. While CARDIA protocols did not include the use of a bronchodilator, the FHS-Offspring cohort (i.e., at examination 9) performed postbronchodilator measurements in those with airflow limitation. Given the epidemiological nature of this study, we will use prebronchodilator measures of lung function. Airflow limitation is defined as prebronchodilator forced expiratory volume in 1 second (FEV_1_)/forced vital capacity (FVC) ratio < 0.70 or less than the lower limit of normal [30].

### Symptoms

Chronic respiratory symptoms include asthma, airway wall thickening, dyspnea, phlegm, emphysema, and bronchiectasis. CT-based mucus plugging is associated with dyspnea, phlegm, cough, and CB.

### Environmental Exposures + Air pollution

Information on FHS participants’ residential address, PM_2.5_ levels, and proximity to a major road in relation to air quality was determined with a geospatial model. Models contained daily air pollutants, weather, and chemical exposures that integrates satellite-based aerosol measurements, Goddard Earth Observing System atmospheric, weather data, and local land data. Machine learning techniques were employed to refine the model based on ground-level measured exposure data to estimate participant daily exposures at high resolution with excellent out-of-sample cross-validation performance. Proximity to major roads was calculated via the U.S. Census Feature Class A1, A2, and A3 data [31–33].

### Marijuana Exposure

Information on CARDIA participants’ marijuana smoking habits was collected from Y0 to Y25. Lifetime marijuana use was defined as the number of joints smoked from Y0 onwards, including Y20 and Y25 [34].

### Cardiorespiratory Fitness

Participants had a symptom-limited, graded treadmill exercise test to measure cardiorespiratory fitness during the Y20 CARDIA exam. The modified Balke protocol consists of nine 2-minute stages of increasing difficulty in both speed and grade to a maximum of 5.6 mph at 25% grade (19.0 metabolic equivalent task). The test duration and maximal metabolic equivalent task were recorded [28, 35].

### Genetics

NIH TOPMed whole-genome sequencing will be used on both cohorts to identify common and rare variants associated with CT mucus plugging [36]. The NIH TOPMed Study performed alignment and variant calling using a standardized pipeline detailed at nhlbiwgs.org, yielding high-quality whole genome sequencing variant calls.

### Hypotheses

There are three primary hypotheses:

♦ Hypothesis 1: CT-based mucus plugging is associated cross-sectionally with lower FEV_1_, higher burden of respiratory symptoms, and CB; persistent mucus plugs over time are associated with a decline in FEV_1_ and future development of COPD.
♦ Hypothesis 2: Air pollution and lifetime marijuana use are associated with increased odds of CT-based mucus plugging, whereas cardiorespiratory fitness is protective against airway mucus plug formation.
♦ Hypothesis 3: Common and rare variants are associated with CT-based mucus plugging.

### Analysis Plan

To evaluate Hypothesis 1, we will conduct separate analyses for FHS and CARDIA. In CARDIA, linear and logistic multivariable models will be used to assess the cross-sectional relationships between CT-based mucus plugging and FEV_1_ % predicted (continuous), cough, phlegm, dyspnea, and bronchitis episodes (all binary). We will consider the following covariates: age, sex, race, center, education level, BMI, ever-smoking status, pack-year, and history of asthma. In FHS, linear mixed-effect models will be used for FEV_1_ and generalized mixed-effect models for binary outcomes: cough, phlegm, dyspnea, and CB. In these models, fixed effects, except for race, will be like those used in CARDIA. Since FHS includes participants with familial relatedness (parents and children), we will include a random effect to account for familial aggregation. Finally, we will conduct sex-stratified analyses for these outcomes, as preliminary studies suggest women have higher odds of mucus plugging.

We will use the CARDIA cohort to assess the association between CT-based mucus plugs and the outcome change in FEV_1_ over time, using Y20 and Y25 CT data. For these longitudinal analyses, we will classify changes in mucus plugging into two groups: Persistent Positive (mucus plugs at both Y20 and Y25) + Newly Formed (mucus plugs at Y25 but not at Y20) vs. Persistent Negative (no mucus plugs at both Y20 and Y25) + Resolved (mucus plugs at Y20 but not at Y25). The two latter groups combined are the referent group. We will use linear mixed models and generalized mixed models to assess whether that binary variable is associated with the outcomes decline in FEV_1_ and COPD at the Y30 exam. Decline in FEV_1_ will be defined as Y30 minus Y20 divided by 10, yielding changes per year. Unfortunately, CARDIA has no spirometry data at Y25. COPD is defined as a pre-bronchodilator FEV_1_/FVC ratio <0.7, an acceptable approach for epidemiologic studies [37, 38]. The cutoff of 0.7 has been validated in population-based studies [39]. Models for FEV_1_ decline will include age, sex, race, center, education level, BMI, ever-smoking status, pack-year, baseline FEV_1_, and history of asthma. Time-variant variables include pack-year, smoking status, and BMI. Also, we will perform analyses to explore sex and race differences in the associations of interest, drawing from the sex and racial-balanced number of CARDIA participants.

### Power calculation and sample size for Hypothesis 1

Detectable effect sizes are calculated for FEV_1_ decline and future COPD to reach at least 80% power, assuming a sample size of 2,600 participants with CARDIA Y20 and Y25 CT data and an alpha value of 0.05. If the prevalence of persistent mucus plugging is 10%, we detect a 53mL FEV_1_ decline and a 1.87 odds ratio of future COPD. With a prevalence of persistent mucus plugging of 20%, we detect a 40 mL FEV_1_ decline and an odds ratio of 1.61 for future COPD.

To evaluate Hypothesis 2, we will perform separate analyses in CARDIA and FHS. For CARDIA, we will use a logistic model to assess the associated between lifetime marijuana smoking and the outcome CT-based mucus plugging (i.e., binary variable defined as mucus plug score 0 vs. >0). Based on prior studies, marijuana will be categorized to indicate high and low consumption. We will consider adjustments for age, race, education attained, BMI, FEV_1_, presence of emphysema on CT, and asthma history. Also, we will build separate models to examine the relationships between tobacco smoking (ever vs. never, pack-years) and mucus plugging and compared them. Finally, we will explore whether tobacco and marijuana smoking have independent or additive effects on mucus plug formation. In separate logistic models, we will test the association between fitness (i.e., treadmill duration) and mucus plugging. Further, we think that fitness may counter the effect of tobacco smoking on mucus formation. Thus, in secondary analyses, we will test whether fitness modifies (i.e., mitigates) the impact of cigarette smoking status and pack-years on CT-based mucus plugging using the appropriate interactions terms. For FHS, a generalized mixed model will test the association between air pollution and distance to roadways (exposures) with CT-based mucus plugging as the outcome. Based on previous studies, we will consider using two exposures: 5-year PM_2.5_ average concentration (2004-2008) before CT scanning (2008-2011) and distance to major roads as categories. We will consider the following covariates: age, sex, education level, BMI, smoking (ever vs. never), pack-years, the value of owner-occupied housing, population density in the census track, FEV_1_, and CT measures of emphysema and airway-wall thickening. A random effect will be used to account for FHS familial aggregation.

### Power calculation and sample size for Hypothesis 2

Detectable effect sizes are also computed on the risk of mucus plugging (binary outcome) for fitness and PM_2.5_ and reach at least 80% power, assuming an alpha level of 0.05 and sample sizes of 2,677 (fitness, CARDIA) and 2,340 (PM_2.5_, FHS). CARDIA and FHS sample sizes were smaller due to missing fitness and PM_2.5_ data. In a prevalence of CT-based mucus plugs of 5%, there is a 1.29 relative risk of mucus plugging per 1 SD decrease in fitness and a 1.31 relative risk per 1 SD increase in PM_2.5_ mcg/m^3^. In a prevalence of CT-based mucus plugs of 20%, there is a 1.15 relative risk of mucus plugging per 1 SD decrease in fitness and a 1.16 relative risk per 1 SD increase in PM_2.5_ mcg/m^3^.

To evaluate Hypothesis 3, we will use NIH TOPMed data for common and rare variants. For common variants, we will examine variants with minor allele counts of above 30 in FHS and CARDIA. We will perform a combined analysis, leveraging the single dataset available for all TOPMed [40]. The main analysis will include all participants and adjust for age, sex, current smoking, pack-years, sequencing center/study, race, familial aggregation, and principal ancestry components in a linear mixed model implemented in SAIGE [41].To balance maximum power with the desire for replication, we will apply a tiered strategy previously used and consider ‘Tier 1’ signals those that reach pre-determined levels (see below) in FHS and replicate in CARDIA, and ‘Tier 2’ signals those that reach significance in the analysis of FHS and CARDIA, and that have nominally significant P values in both studies.

Given the importance of cigarette smoking and potential air pollution in mucus plug formation, we will also perform secondary analysis using common variants including both the main effect and a gene by environment interaction using a two-degree of freedom joint test that improves power in the setting of interaction [42]. Due to the differences in prevalence among races, we will also examine results by ancestry and perform admixture analysis in Black participants in CARDIA using LAMP-LD, as we have previously performed [43].

For rare variants, aggregating variants can increase power. We will use the recently developed ACAT-O (aggregated Cauchy association test, omnibus), which combines set-based tests including burden, SKAT, and ACAT-V, improves power in the setting of sparsity, and is superior to earlier methods such as SKAT-O [44]. We will test aggregating by gene or by sliding window region and also examine the effects of limiting our analysis to coding variants (nonsynonymous, stop, or splice) or those with putative regulatory function (e.g., based on overlap with open chromatin in lung-related cells) [45].

We also anticipate examining a specific set of ∼100 genes known to be important for mucus dysfunction (e.g., TP53) and muco-obstructive diseases such as COPD and bronchiectasis (e.g., *FAM13A*, *MUC5B*, *MUC5AC*, *CFTR*, and *DNAH5)*. This secondary analysis decreases the multiple comparison penalty from genome-wide approaches while enabling a more detailed examination of genes with a higher likelihood of association. As a secondary analysis, we will perform a genome-wide association.

We will analyze heritability, genetic correlation, functional enrichment, and potential causal genes to analyze and interpret our genetic association results. We will use GCTA-SC and GCTA-LDMS in our European sample to estimate heritability and genetic correlation between this CT phenotype and lung function [46]. To identify potential causal genes, we will rely on multiple approaches, including expression quantitative trait loci (eQTL) in lung and blood, examination of DNAase hypersensitivity sites, chromosome conformation (Hi-C), and gene set and pathway methods (DEPICT), as previously performed [47].

### Power calculation and sample size for Hypothesis 3

When assessing genetic variance to CT-based airway mucus plugs, in a total sample size of 5,427 (FHS + Y20 CARDIA) and assuming a prevalence of mucus plugging of 10%, we have ∼ 80% power to identify a variant at a P < 1 × 10^-4^ with an allele frequency of 40% and an odds ratio of ∼1.3. In addition, we have > 80% power at alpha = 5 × 10^-8^ in our genome-wide analysis to identify a variant that increases risk by 1.45.

### Ethics and Dissemination

The present study was approved by the Mass General Brigham Institutional Review Board (2022P000723). Participants were recruited voluntarily for these studies. Written informed consent was obtained for CT scanning, clinical data, blood sample collection, and genomic analyses. Participants were under no obligation to participate. A potential risk is the risk of participant de-identification or the release of personal health information. We will use genotype and phenotype files, which are only identified by study ID numbers; the investigators are not members of the Data Coordinating Center of these studies and will therefore not have access to any information that is not already de-identified. To protect against inadvertent disclosure of identity, we will make all study data available to study investigators using the de-identified study identifications assigned by the CARDIA and FHS studies. The links to identifiers are not available to the investigators. All study data is stored on a secured, password-protected computer system at Brigham and Women’s Hospital (BWH) and CARDIA and FHS Data Coordinating Centers.

We plan to disseminate the findings of this study through peer-reviewed journals and professional scientific conferences, such as the American Thoracic Society’s annual Conference.

## Data Availability

All data produced in the present study are available upon reasonable request to the authors.

## Footnotes

### Author Contributions

Guarantors of the integrity of the study, A.A.D.; study concepts/study design or data analysis/interpretation, A.A.D, R.E., M.C., R.K., M.R.; obtained funding, A.A.D., M.C.; data acquisition, M.R., G.O.,N.A., A.Y., M.H., M.A., R.E., A.A.D.; manuscript drafting or manuscript revision for important intellectual content, all authors; drafted the article and critically revised it for intellectual content, A.A.D., M.A.; approval of the final version of the submitted manuscript, all authors; agree to ensure any questions related to the work are appropriately resolved, all authors.

### Funding

This work was supported by the National Heart Lung and Blood Institute (Grant R01-HL164824). The Coronary Artery Risk Development in Young Adults Study (CARDIA) is supported by contracts 75N92023D00002, 75N92023D00003, 75N92023D00004, 75N92023D00005, and 75N92023D00006 from the National Heart, Lung, and Blood Institute (NHLBI). The funders of this project were not involved in the writing, editing, or decision to publish this study. This manuscript was reviewed and approved by the CARDIA Publications and Presentations Committee.

### Conflict of Interests

Maya Abdalla and Rim Elalami have no conflict of interests to disclose. Dr Cho reported receiving grants from Bayer. Dr. O’Connor reports no conflict of interests. Dr Rice reported receiving research grant funding from the NIH and receiving expert testimony fees from the Conservation Law Foundation. Dr. Horowitz reported consulting fees from Bayer and is a member of GE medical imaging advisory board. Dr. Akhoundi had no conflicts of interest to disclose. Dr. Yen reported receiving salary support. Dr Kalhan reported grants from AstraZeneca, PneumRx/BTG and Spiration to Northwestern University; consulting fees from CVS Caremark, AstraZeneca, GlaxoSmithKline, and CSA Medical; payment from GlaxoSmithKline, AstraZeneca, and Boehringer Ingelheim and is a member of CSA Medical advisory board. Dr Diaz reported receiving personal fees from Boehringer Ingelheim and having a patent for Methods and Compositions Relating to Airway Dysfunction pending (701586-190200USPT).

## Notes

### Author Declarations

Mass General Brigham Institutional Review Board gave ethical approval for this work (2022P000723). This work was supported by the National Heart Lung and Blood Institute (Grant R01-HL164824).

## Reference

1. Boucher, R.C., Muco-obstructive lung diseases. Reply. The New England journal of medicine, 2019. 381(10): p. e20.

2. Fahy, J.V. and B.F. Dickey, Airway mucus function and dysfunction. New England journal of medicine, 2010. 363(23): p. 2233–2247.

3. Svenningsen, S., et al., CT and functional MRI to evaluate airway mucus in severe asthma. Chest, 2019. 155(6): p. 1178–1189.

4. Dunican, E.M., et al., Mucus plugs in patients with asthma linked to eosinophilia and airflow obstruction. The Journal of clinical investigation, 2018. 128(3): p. 997–1009.

5. Chassagnon, G. and P.-R. Burgel, Mucus Plugs in Medium-sized Airways: A Novel Imaging Biomarker for Phenotyping Chronic Obstructive Pulmonary Disease. 2021, American Thoracic Society. p. 932–934.

6. Okajima, Y., et al., Luminal Plugging on Chest CT Scan: Association With Lung Function, Quality of Life, and COPD Clinical Phenotypes. Chest, 2020. 158(1): p. 121–130.

7. Dunican, E.M., et al., Mucus Plugs and Emphysema in the Pathophysiology of Airflow Obstruction and Hypoxemia in Smokers. Am J Respir Crit Care Med, 2021. 203(8): p. 957–968.

8. Kim, V., et al., Mucus plugging on computed tomography and chronic bronchitis in chronic obstructive pulmonary disease. Respir Res, 2021. 22(1): p. 110.

9. Mettler, S.K., et al., Silent airway mucus plugs in COPD and clinical implications. Chest, 2023.

10. Ford, E.S., et al., Trends in the prevalence of obstructive and restrictive lung function among adults in the United States: findings from the National Health and Nutrition Examination surveys from 1988-1994 to 2007-2010. Chest, 2013. 143(5): p. 1395–1406.

11. Ma, J., et al., Temporal trends in mortality in the United States, 1969-2013. Jama, 2015. 314(16): p. 1731–1739.

12. Button, B., et al., Roles of mucus adhesion and cohesion in cough clearance. Proceedings of the National Academy of Sciences, 2018. 115(49): p. 12501–12506.

13. Kesimer, M., et al., Airway mucin concentration as a marker of chronic bronchitis. New England Journal of Medicine, 2017. 377(10): p. 911–922.

14. Çolak, Y., et al., Prognostic significance of chronic respiratory symptoms in individuals with normal spirometry. European Respiratory Journal, 2019. 54(3).

15. Allinson, J.P., et al., The presence of chronic mucus hypersecretion across adult life in relation to chronic obstructive pulmonary disease development. American journal of respiratory and critical care medicine, 2016. 193(6): p. 662–672.

16. Guerra, S., et al., Chronic bronchitis before age 50 years predicts incident airflow limitation and mortality risk. Thorax, 2009. 64(10): p. 894–900.

17. Puhan, M.A., Chronic respiratory symptoms but normal lung function: substantial disease burden but little evidence to inform practice. 2019, Eur Respiratory Soc.

18. Mejza, F., et al., Prevalence and burden of chronic bronchitis symptoms: results from the BOLD study. European Respiratory Journal, 2017. 50(5).

19. Woodruff, P.G., et al., Clinical significance of symptoms in smokers with preserved pulmonary function. New England Journal of Medicine, 2016. 374(19): p. 1811–1821.

20. Hasselgren, M., et al., Estimated prevalences of respiratory symptoms, asthma and chronic obstructive pulmonary disease related to detection rate in primary health care. Scandinavian journal of primary health care, 2001. 19(1): p. 54–57.

21. Diaz, A.A., et al., Airway-Occluding Mucus Plugs and Mortality in Patients With Chronic Obstructive Pulmonary Disease. JAMA, 2023. 329(21): p. 1832–1839.

22. Liu, G.Y. and R. Kalhan, Impaired respiratory health and life course transitions from health to chronic lung disease. Chest, 2021. 160(3): p. 879–889.

23. Silverman, E.K., et al., Genetic epidemiology of severe, early-onset chronic obstructive pulmonary disease: risk to relatives for airflow obstruction and chronic bronchitis. American journal of respiratory and critical care medicine, 1998. 157(6): p. 1770–1778.

24. Chand, H.S., et al., A genetic variant of p53 restricts the mucous secretory phenotype by regulating SPDEF and Bcl-2 expression. Nat Commun, 2014. 5: p. 5567.

25. Friedman, G.D., et al., CARDIA: study design, recruitment, and some characteristics of the examined subjects. Journal of clinical epidemiology, 1988. 41(11): p. 1105–1116.

26. Kannel, W.B., et al., An investigation of coronary heart disease in families: the Framingham Offspring Study. American journal of epidemiology, 1979. 110(3): p. 281–290.

27. Splansky, G.L., et al., The third generation cohort of the National Heart, Lung, and Blood Institute’s Framingham Heart Study: design, recruitment, and initial examination. American journal of epidemiology, 2007. 165(11): p. 1328–1335.

28. Diaz, A.A., et al., Association between cardiorespiratory fitness and bronchiectasis at CT: A long-term population-based study of healthy young adults aged 18–30 years in the CARDIA study. Radiology, 2021. 300(1): p. 190–196.

29. Rice, M.B., et al., Exposure to traffic emissions and fine particulate matter and computed tomography measures of the lung and airways. Epidemiology, 2018. 29(3): p. 333–341.

30. Oelsner, E.C., et al., Harmonization of respiratory data from 9 US population-based cohorts: the NHLBI Pooled Cohorts Study. American journal of epidemiology, 2018. 187(11): p. 2265–2278.

31. Rice, M.B., et al., Long-term exposure to traffic emissions and fine particulate matter and lung function decline in the Framingham heart study. American journal of respiratory and critical care medicine, 2015. 191(6): p. 656–664.

32. Synn, A.J., et al., Ambient air pollution exposure and radiographic pulmonary vascular volumes. Environmental Epidemiology, 2021. 5(2).

33. Rice, M.B., et al., Ambient air pollution exposure and risk and progression of interstitial lung abnormalities: the Framingham Heart Study. Thorax, 2019. 74(11): p. 1063–1069.

34. Pletcher, M.J., et al., Association between marijuana exposure and pulmonary function over 20 years. JAMA, 2012. 307(2): p. 173–81.

35. Benck, L.R., et al., Association between cardiorespiratory fitness and lung health from young adulthood to middle age. American journal of respiratory and critical care medicine, 2017. 195(9): p. 1236–1243.

36. Prokopenko, D., et al., Whole-genome sequencing in severe chronic obstructive pulmonary disease. American journal of respiratory cell and molecular biology, 2018. 59(5): p. 614–622.

37. Khalid, F., et al., Prevalence and Population Attributable Risk for Early Chronic Obstructive Pulmonary Disease in U.S. Hispanic/Latino Individuals. Ann Am Thorac Soc, 2022. 19(3): p. 363–371.

38. Colak, Y., et al., Prevalence, Characteristics, and Prognosis of Early Chronic Obstructive Pulmonary Disease. The Copenhagen General Population Study. Am J Respir Crit Care Med, 2020. 201(6): p. 671–680.

39. Bhatt, S.P., et al., Discriminative Accuracy of FEV1:FVC Thresholds for COPD-Related Hospitalization and Mortality. JAMA, 2019. 321(24): p. 2438–2447.

40. Hobbs, B.D., et al., Genetic loci associated with chronic obstructive pulmonary disease overlap with loci for lung function and pulmonary fibrosis. Nat Genet, 2017. 49(3): p. 426–432.

41. Zhou, W., et al., Efficiently controlling for case-control imbalance and sample relatedness in large-scale genetic association studies. Nat Genet, 2018. 50(9): p. 1335–1341.

42. Manning, A.K., et al., Meta-analysis of gene-environment interaction: joint estimation of SNP and SNP x environment regression coefficients. Genet Epidemiol, 2011. 35(1): p. 11–8.

43. Parker, M.M., et al., Admixture mapping identifies a quantitative trait locus associated with FEV1/FVC in the COPDGene Study. Genet Epidemiol, 2014. 38(7): p. 652–9.

44. Liu, Y., et al., ACAT: A Fast and Powerful p Value Combination Method for Rare-Variant Analysis in Sequencing Studies. Am J Hum Genet, 2019. 104(3): p. 410–421.

45. Prokopenko, D., et al., Whole-Genome Sequencing in Severe Chronic Obstructive Pulmonary Disease. Am J Respir Cell Mol Biol, 2018. 59(5): p. 614–622.

46. Yang, J., et al., Genetic variance estimation with imputed variants finds negligible missing heritability for human height and body mass index. Nat Genet, 2015. 47(10): p. 1114–20.

47. Sakornsakolpat, P., et al., Genetic landscape of chronic obstructive pulmonary disease identifies heterogeneous cell-type and phenotype associations. Nature genetics, 2019. 51(3): p. 494–505.

